# Associations Between Polygenic Scores for Cognitive and Non-cognitive Factors of Educational Attainment and Measures of Behavior, Psychopathology, and Neuroimaging in the Adolescent Brain Cognitive Development Study

**DOI:** 10.1101/2023.10.27.23297675

**Authors:** Aaron J. Gorelik, Sarah E. Paul, Alex P. Miller, David A.A. Baranger, Shuyu Lin, Wei Zhang, Nourhan M. Elsayed, Hailey Modi, Pooja Addala, Janine Bijsterbosch, Deanna M. Barch, Nicole R. Karcher, Alexander S. Hatoum, Arpana Agrawal, Ryan Bogdan, Emma C. Johnson

## Abstract

**Background:** Both cognitive and non-cognitive (e.g., traits like curiosity) factors are critical for social and emotional functioning and independently predict educational attainment. These factors are heritable and genetically correlated with a range of health-relevant traits and behaviors in adulthood (e.g., risk-taking, psychopathology). However, whether these associations are present during adolescence, and to what extent these relationships diverge, could have implications for adolescent health and well-being.

**Methods:** Using data from 5,517 youth of European ancestry from the ongoing Adolescent Brain Cognitive Development^SM^ Study, we examined associations between polygenic scores (PGS) for cognitive and non-cognitive factors and outcomes related to cognition, socioeconomic status, risk tolerance and decision-making, substance initiation, psychopathology, and brain structure.

**Results:** Cognitive and non-cognitive PGSs were both positively associated with cognitive performance and family income, and negatively associated with ADHD and severity of psychotic-like experiences. The cognitive PGS was also associated with greater risk-taking, delayed discounting, and anorexia, as well as lower likelihood of nicotine initiation. The cognitive PGS was further associated with cognition scores and anorexia in *within-sibling* analyses, suggesting these results do not solely reflect the effects of assortative mating or passive gene-environment correlations. The cognitive PGS showed significantly stronger associations with cortical volumes than the non-cognitive PGS and was associated with right hemisphere caudal anterior cingulate and pars-orbitalis in *within-sibling* analyses, while the non-cognitive PGS showed stronger associations with white matter fractional anisotropy and a significant *within-sibling* association for right superior corticostriate-frontal cortex.

**Conclusions:** Our findings suggest that PGSs for cognitive and non-cognitive factors show similar associations with cognition and socioeconomic status as well as other psychosocial outcomes, but distinct associations with regional neural phenotypes in this adolescent sample.

## INTRODUCTION

As educational attainment (EduA) is among the strongest predictors of positive outcomes across the lifespan (e.g., income, health, well-being; Gutacker et al., 2023; Raghupathi & Raghupathi, 2020; Zajacova & Lawrence, 2018), it is important to understand what contributes to educational attainment. Despite often being thought of as a purely “environmental” factor, EduA itself is moderately heritable (h^2^=0.41-0.47; Heath et al., 1985; Silventoinen et al., 2020), and the genetic contributions to EduA can be broken down into component traits. Evidence that EduA is strongly impacted by cognitive ability and a set of broadly defined “non-cognitive skills” (e.g., emotion regulation and personality traits such as grit and curiosity; Chamorro-Premuzic & Furnham, 2003; Duckworth et al., 2007, 2019; Kovas et al., 2015; Malanchini et al., 2019; Noftle & Robins, 2007) has inspired recent approaches that have deconstructed the genetic architecture of EduA into cognitive and non-cognitive components that have shared and unique associations with EduA-related phenotypes (e.g., risk-taking and psychopathology; Malanchini et al., 2023; Tucker-Drob et al., 2016; Tucker-Drob & Harden, 2012). The extent to which these differential associations extend to childhood, before education has been completed, remains poorly understood (Malanchini et al., 2023). In the present study, we examined whether cognitive and non-cognitive polygenic score (PGS) associations mirror those found in primarily adult sample studies and use *within-sibling* analyses to assess whether results show evidence of confounding (e.g., by population stratification, assortative mating, or passive gene-environment correlations).

### Deconstructing the Genetic Architecture of Educational Attainment into Cognitive and Non-Cognitive Components

Demange et al., 2021 demonstrated that EduA can be genetically parsed into cognitive and non-cognitive factors. To study genetic influences on “non-cognitive skills,” Demange et al. 2021 performed a novel “GWAS-by-subtraction” by residualizing the genetic effects of EduA (N=1,131,881; Lee et al., 2018) on cognitive performance in a Cholesky decomposition using genomic structural equation modeling (Grotzinger et al., 2019), leaving a residual “non-cognitive” genetic factor (N=510,715) and “cognitive” genetic factor (N=257,841; Lee et al., 2018) (see **Figure 1** for overview). In this way, the authors generated a new genome-wide association study (GWAS) of “non-cognitive skills’’ that represents genetic influences on EduA that are *not* shared with cognitive performance (by design, the “non-cognitive” factor is orthogonal to the “cognitive” factor). Their study found 157 independent loci associated with the non-cognitive factor and observed that the non-cognitive factor showed distinct associations with other relevant phenotypes, including positive genetic correlations with risk tolerance and some psychiatric disorders (e.g., bipolar disorder and schizophrenia), and was positively genetically correlated with personality traits including conscientiousness, extraversion, and agreeableness while the cognitive factor showed negative or null correlations with these same phenotypes (Demange et al., 2021).

**Figure 1.**
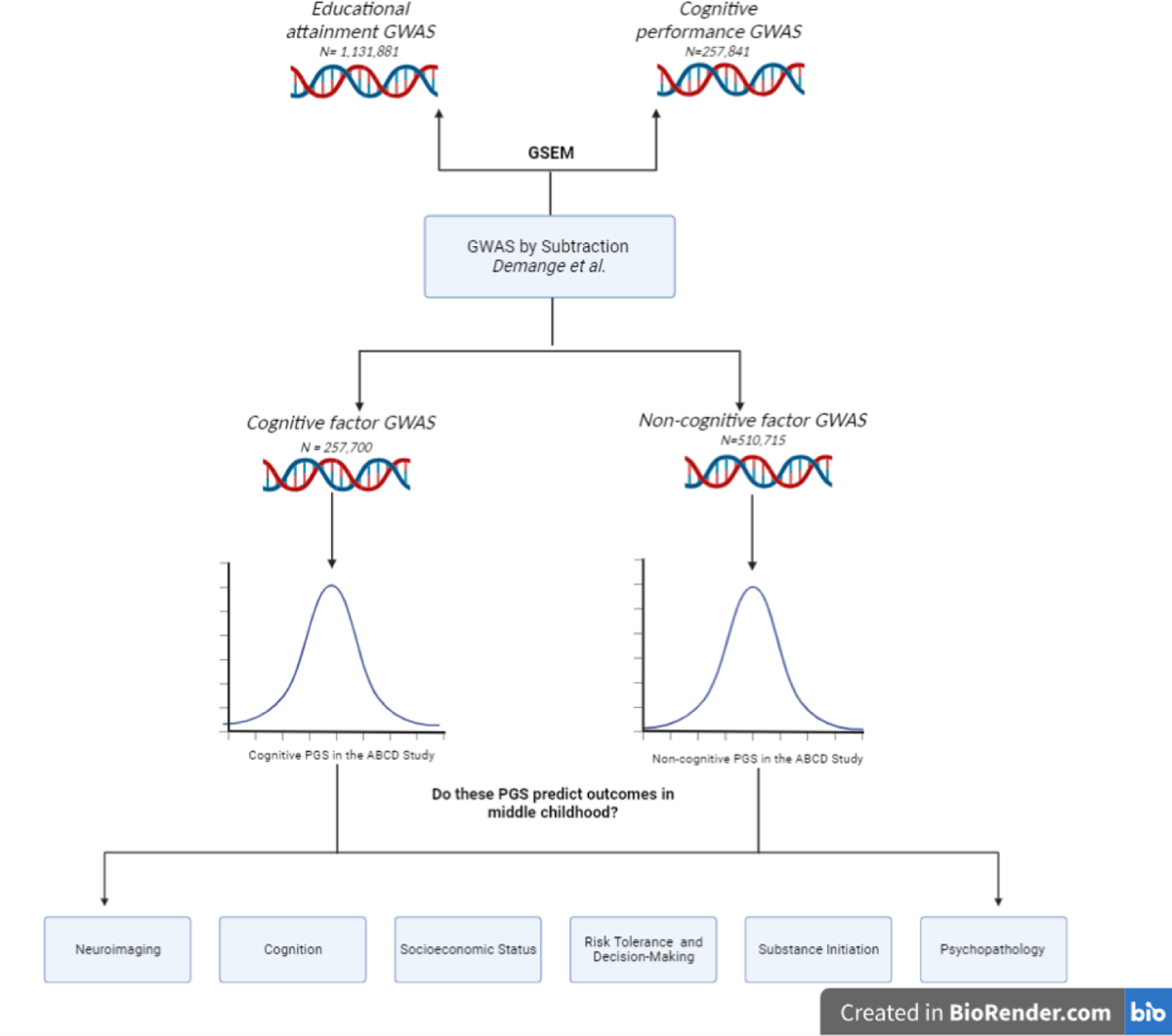
Overview of study design. Cognitive and non-cognitive PGSs were created using summary statistics from a GWAS-by-subtraction by Demange *et al.,* 2021. We then tested whether these PGSs were associated with cognition, socioeconomic status, risk tolerance and decision-making, substance initiation, psychopathology, and neuroimaging phenotypes in the Adolescent Brain Cognitive Development (ABCD) Study.

While the GWASs mentioned above consisted largely of adult participants, adolescence is a critical stage of both cognitive and non-cognitive development. Prior studies have shown that early differences in cognitive and non-cognitive factors (e.g., personality; Chamorro-Premuzic & Furnham, 2003; Noftle & Robins, 2007) may contribute to EduA, employment outcomes, and overall success later in life (Duckworth et al., 2019; Moffitt et al., 2011). A recent preprint reported that the association between a non-cognitive PGS and academic achievement nearly doubled between the ages of 7 and 16 (Malanchini et al., 2023), suggesting that genetic factors related to non-cognitive facets of EduA may become particularly influential during adolescence, a critical developmental period for factors related to EduA. Importantly, examinations of genetic associations between cognitive and non-cognitive factors and brain imaging have mostly been conducted in adult samples (Demange et al., 2021). Little is known of the brain mechanisms related to these factors in adolescence, a critical time period for neural plasticity. In sum, despite adolescence being a critical time for future academic achievement, little is known about the cognitive and non-cognitive influences on EduA and their contributions to relevant traits, behaviors, and brain structure in adolescence.

### The Current Study

In the current study, we estimated the associations between PGS for cognitive and non-cognitive factors of EduA and outcome measures among children of European ancestry enrolled in the ongoing Adolescent Brain Cognitive Development (ABCD) Study (Volkow et al., 2018; analytic n up to 5,517). We focused on behavioral phenotypes implicated in studies on adults (i.e., cognition, family socioeconomic status, risk tolerance & decision-making, substance initiation, psychopathology; Lee et al., 2018) and also examined associations with brain structure. Finally, as prior research has shown that genetic confounds (e.g., assortative mating; Horwitz et al., 2023) can inflate GWAS test statistics and polygenic score associations (Okbay et al., 2022), we performed *post hoc within-sibling* analyses to assess whether any significant associations may be independent of assortative mating, passive gene-environment correlation, or other sociodemographic confounders.

## METHODS

### Participants

The ongoing Adolescent Brain and Cognitive Development ^SM^ (ABCD) Study^®^ is a longitudinal study following 11,879 children (ages 8.9-11 at baseline; born between 2005-2009) recruited from 22 research sites across the United States to study the development of complex behavior and biology from middle childhood to late adolescence/young adulthood in the context of experience and genetic background (Volkow et al., 2018). It includes a family-based component in which twin (n=2108), triplet (n=30), non-twin siblings (n=1,589), and singletons (n=8,148) were recruited. Caregivers provided written informed consent and their children provided verbal assent. For the present study, we used data from the baseline (2016-2018; ages: 9-11) as well as 2-year (FU2; 2018-2021; ages 11-13) and 3-year (FU3; 2021-2022; ages 9-14)) follow-up sessions. Analyses were only conducted in individuals with genetic ancestry most similar to those of European genetic ancestry reference populations (see **Genetic Data section** below), due to the lack of relevant well-powered discovery GWAS in other ancestries and the low predictive utility of PGS when applied across ancestries (Martin et al., 2019). After excluding individuals with missing outcome or covariate data, described below, analytic Ns ranged from 5,168-5,517 (baseline), 3352-5,006 (FU2), and 2,968-4745 (FU3), with the exception of substance use data as described below.

### Measures

Cognitive performance, risk tolerance/decision making, substance use initiation, psychopathology, and neuroimaging data were drawn from the baseline, FU2, and FU3 assessments from the National Institute of Mental Health Data Archive (NDA; https://nda.nih.gov/); data release 4.0 and 5.0). Socioeconomic status and genomic data (release 3.0) were derived from the baseline session.

#### Cognition

Crystallized and fluid intelligence as well as their total composite were estimated from the NIH ToolBox assessment (Luciana et al., 2018).

#### Socioeconomic status

Caregiver-reported combined past-12-month family income (Luciana et al., 2018) and neighborhood deprivation index, a composite of neighborhood socioeconomics, were drawn from baseline data (Fan et al., 2021).

#### Risk tolerance and decision-making

Risk tolerance was measured at baseline using the single item, “I enjoy taking risks” from the sensation seeking scale of the UPPS-P (Watts et al., 2020). Delayed discounting was measured at baseline using the single item cash choice task where youth decide whether they would “rather have $75 in three days or $115 in 3 months” (Luciana et al., 2018; Wulfert et al., 2002). This single item measure was used, as opposed to behavioral data acquired (Kohler et al., 2022) due to quality control procedures of these data resulting in the exclusion of large amounts of baseline data.

#### Substanc use initation

Lifetime alcohol, nicotine, and cannabis initiation was assessed from the annual substance use interview and mid-year substance phone interviews (Miller et al., 2023; Volkow et al., 2018) from baseline to FU3. Individuals endorsing substance use only in the context of religious ceremonies were excluded. Substance naive participants endorsed no substance initiation (e,g., substance naive individuals being compared to those who initiatiate cannabis use had not used alcohol, tobacco or other substances). Substance initiation analytic Ns ranged from 2,968 to 4,745 **(see Supplemental Table 1-2 for additional information).**

#### Psychopathology

We assessed seven mental health related measures. Given that schizophrenia typically presents later in adolescence, we included a summary score of the severity of youth psychotic-like experiences (Karcher & Barch, 2021) from FU2. We also included a caregiver-reported diagnosis screener for autism from the baseline assessment (Barch et al., 2018). In a similar vein, as most psychopathologies assessed tend to onset in mid-adolescence, when possible we used baseline to FU2 to generate lifetime KSADS-5 diagnoses (Kaufman et al., 1997) to capture this critical time period for all assessed mental health diagnoses. Lifetime KSADS-5 diagnoses were created for: obsessive compulsive disorder (OCD), anorexia, bipolar disorder, major depressive disorder (MDD), and attention deficit hyperactivity disorder (ADHD). The analytical N for these measures ranged from 3,352 to 3,737 and all definitions and items used to create the lifetime mental health diagnoses measures can be found in **Supplemental Table 1.**

#### Neuroimaging

Indices of gray matter structure (i.e., cortical thickness, cortical surface area, and subcortical and cortical volumes) and white matter tracts (i.e., fractional anisotropy [FA], mean diffusivity [MD] were derived using the Desikan-Killiany atlas (Desikan et al., 2006) and Atlas Tract (Basser et al., 1994), respectively. No task-related functional magnetic resonance imaging (fMRI) data were examined due to test–retest reliability concerns (Elliott et al., 2020). Acquisition and preprocessing methodology (Casey et al., 2018; Hagler et al., 2019) as well as additional information can be found in the Supplemental Table 3 and Supplemental Methods.

### Polygenic Scores

We used PRS-CS (Ge et al., 2019) to calculate polygenic scores in the European ancestry subset of the ABCD Study sample, using effect sizes from the Demange et al. GWAS of “cognitive skills” (GWAS catalog accession GCST90011875; effective n=257,700; Demange et al., 2021) and “non-cognitive skills” (GWAS catalog accession GCST90011874; effective n=510,795; Demange et al., 2021) and the 1000 Genomes Phase 3 European reference panel (1000 Genomes Project Consortium et al., 2015). We used the ‘auto’ function of PRS-CS, allowing the software to learn the global shrinkage parameter from the data **(see Supplemental Methods for details)**.

### Statistical Analyses

Analyses were pre-registered on the Open Science Framework and conducted using mixed effects models implemented using lmer (for continuous outcomes) and glmer (for dichotomous outcomes) from the lme4 package (Austin, 2010; Bates et al., 2022) in R (v4.3; R Core Team). Age, sex, and the first 10 genetic principal components were included as fixed effect covariates with family ID and recruitment site as random intercepts to account for data dependence. For imaging models, recruitment site was replaced by MRI serial number. Imaging models also included MRI manufacturer, global brain metrics representing the mean for each modality, and mean motion for DTI as fixed effects. We used false discovery rate (FDR; Benjamini & Hochberg, 1995) to account for multiple testing (p_fdr_<0.05); FDR was applied separately to the non-imaging phenotypes and each respective imaging modality.

We deviated from our pre-registered analyses in two ways: First, we tested whether the regression coefficients for the cognitive and non-cognitive PGS significantly differed from each other in each model (p_diff_; **Supplemental Methods).** Second, we conducted *post hoc within-sibling* analyses to assess whether any significant associations arising from primary analyses (aside from familial SES due to sibling similarity) may plausibly represent direct genetic effects. Significant *within-sibling* effects indicate that these associations are unconfounded by population stratification, assortative mating, passive gene-environment correlations, and other potential population-level confounds (though it should be noted that active and evocative rGE will still influence within-sibling variation in PGS effects; Brumpton et al., 2020; Howe et al., 2022; Young et al., 2018). For these analyses, we included both the family mean PGS and a sibling’s deviation from their family mean PGS as predictors in a mixed-effect model, as has been done previously (Selzam et al., 2019) **(see Supplemental Methods for analysis details)**.

## RESULTS

Demographic descriptive statistics for the baseline analytic sample (max N=5,517) are available in **Table 1**. The cognitive and non-cognitive polygenic scores (PGS) were negatively correlated with one another (r=-0.12, p<2e-16).

**Table 1:** ABCD European Ancestry Baseline Demographic Table.

| Variable | Mean (SD)/n (%) |
| --- | --- |
|  | **baseline** |
| *Sex (male)* | 2612 (47.0%) |
| *Age (years)* | 9.93(0.63) |
| *Household income* |  |
| <\$35,000 | 375(7.1%) |
| \$35,000-\$49,000 | 283(5.1%) |
| \$50,000-\$74,999 | 717(13.5%) |
| \$75,000-\$99,999 | 896(16.9%) |
| \$100,000-\$199,999 | 2178(41.1%) |
| \$200,000 | 849(16.0%) |
| *Highest caregiver education* |  |
| Less than high school | 25 (0.45%) |
| High school degree or equivalent | 188(3.4%) |
| Some college, associate degree | 1046(18.8%) |
| College degree | 1753(31.6%) |
| Master's degree | 1723(31.0%) |
| Doctorate/professional degree | 829(14.8%) |
| *Parental marital status* |  |
| Married or co-habiting | 4721(85.1%) |
| Widowed | 41(0.7%) |
| Divorced/separated | 620(11.2%) |
| Never married | 164(3.0%) |

### Behavioral and sociodemographic outcomes

#### Cognition (Figure 2A; Supplemental Table 4)

The cognitive PGS was positively associated with all 3 cognition measures (i.e., fluid, crystallized, total score); Bs>0.136, p_fdrs_<3.51E-24). The non-cognitive PGS was associated with crystallized intelligence and total scores (Bs>0.05, p_fdrs_<7.64E-04), but not fluid intelligence (B=0.013, p_fdr_=0.715). For all 3 measures, the cognition scores were more strongly associated with the cognitive PGS relative to non-cognitive PGS (p_diff_<0.05).

**Figure 2:**
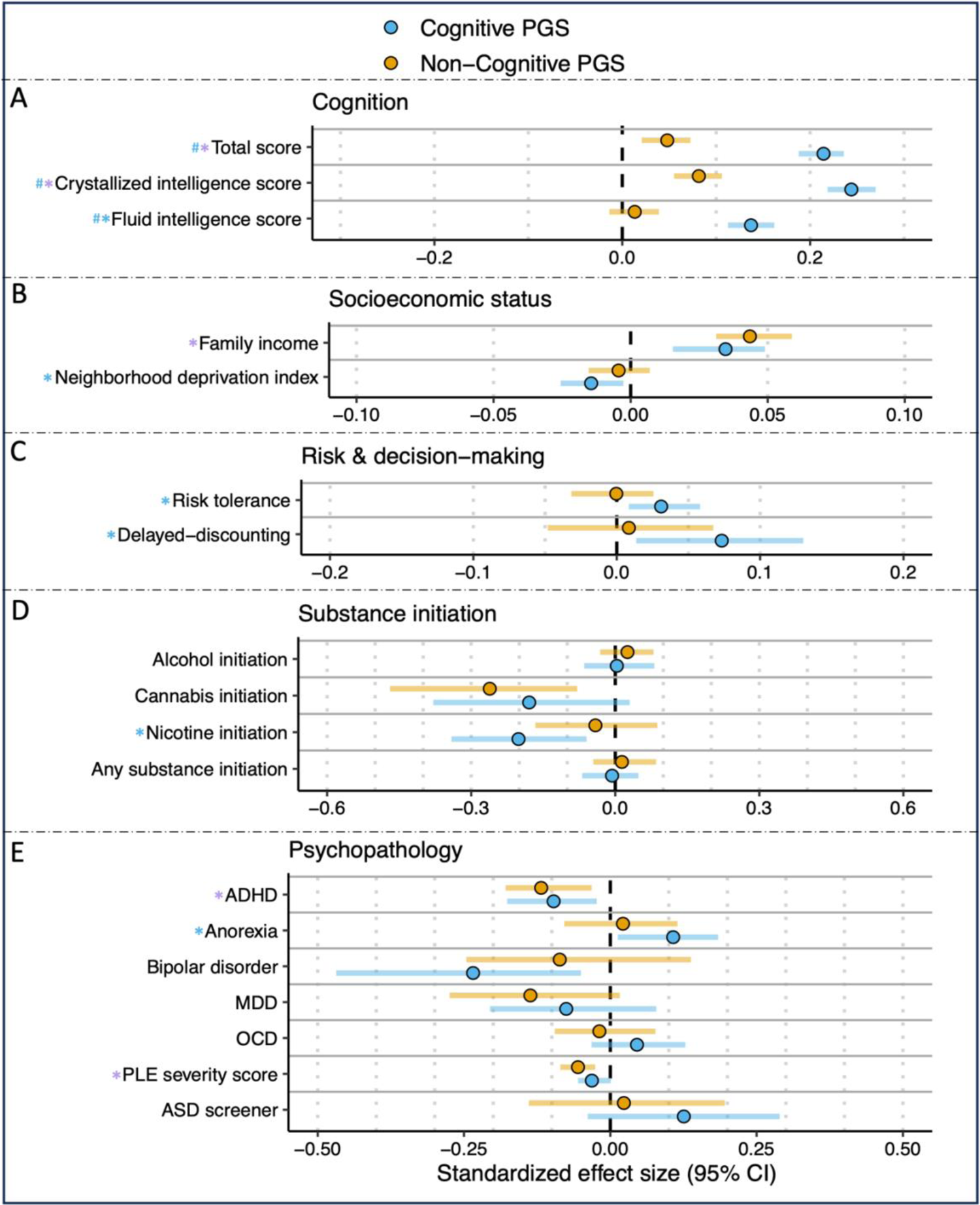
Associations Between Cognitive and Non-cognitive PGS and Neurocognition, SES, Risk-taking and Decision Making, Substance Initiation, and Psychopathology. Blue and purple asterisks correspond to significant associations (p_fdr_<0.05) between the outcome measures of A) cogintion, B) socioeconmic status, C) risk & decision making, D) substance iniation, and E) psychopathology and cognitive PGS or both PGS, respectively. Blue hashtags correspond to associations that are significantly different for the cognitive PGS compared to the non-cognitive PGS. ADHD=attention deficit hyperactivity disorder; ASD=autism spectrum disorder; MDD=major depressive disorder; OCD=obsessive compulsive disorder; PLE=psychotic-like experiences.

**Figure 3:**
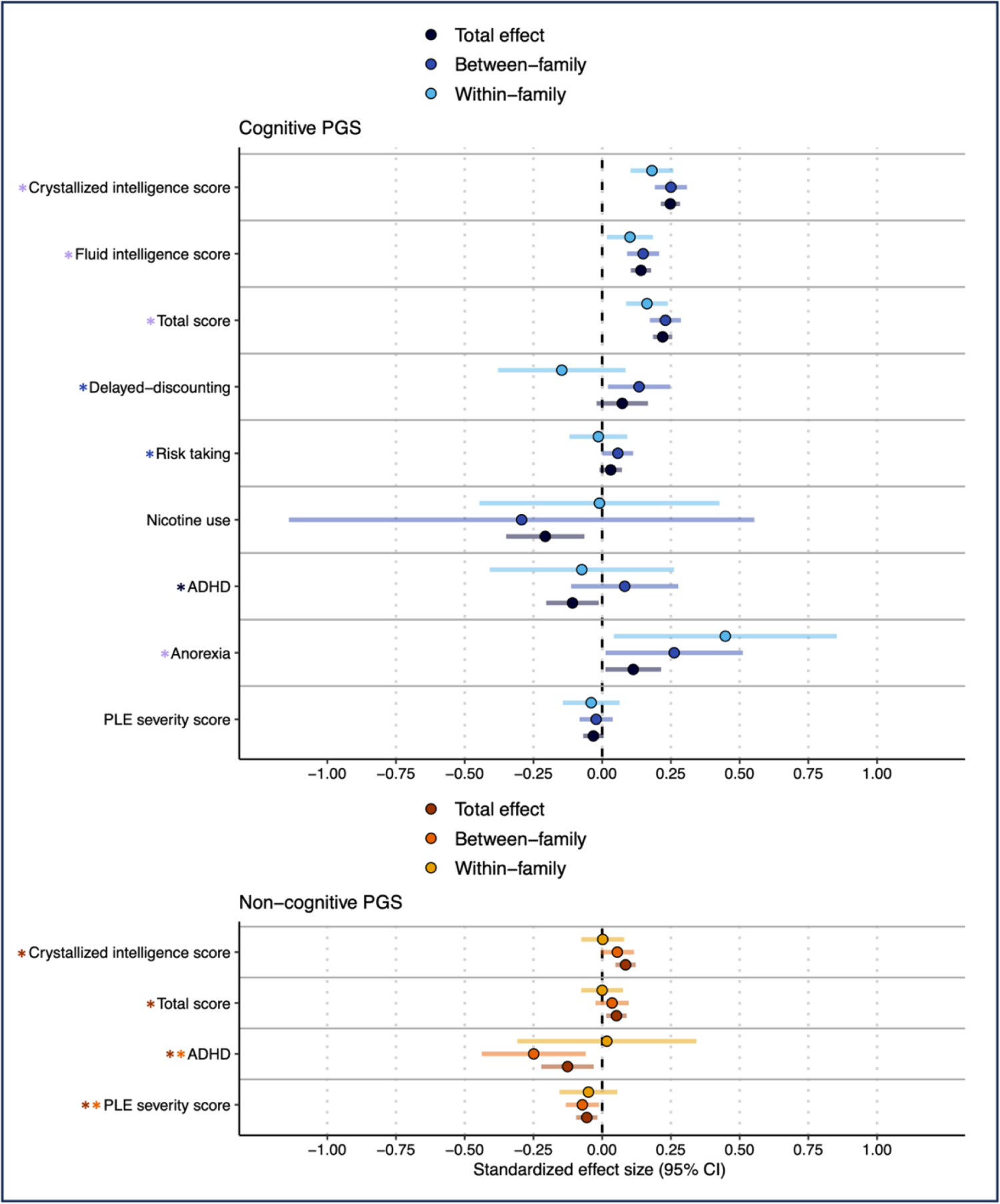
Total, Between, and Within-family Estimates for the Associations Between Cognitive and Non-cognitive PGS and Psychosocial Measures. Total, Within- and between-family associations between Cognitive and Noncognitive PGS (p <0.05) and significant measures in the domains of cognition, socioeconomic status, risk & decision making, and psychopathology (i.e., outcomes with p_fdr_&0.05 in **Figure 2 and Supplemental Table 4**). For the cognitive PGS, black, dark blue, light blue, and purple asterisks correspond to significant total, between-, within-family, and all three associations, respectively. For the non-cognitive PGS, red, orange, and yellow asterisks correspond to significant total, between-, and within-family associations, respectively.

#### SES (Figure 2B; Supplemental Table 4)

The cognitive PGS was associated with greater family SES (family income: B=0.035, p_fdr_=3.36E-05; neighborhood deprivation index: B=-0.014, p_fdr_=0.028). In contrast, the non-cognitive PGS was significantly associated only with total past-12-month combined family income (B =0.04, p_fdr_=3.21E-07), but not the neighborhood deprivation index (B=-0.004, p_fdr_=0.812). Associations with income and neighborhood deprivation did not significantly differ between the cognitive and non-cognitive PGS.

### Brain Structure

#### Global Gray Matter Indices (Figure 4A**; Supplemental Table 6).**

Both cognitive and non-cognitive PGS were positively associated with all (n=9) global volumes of interest including whole brain, whole brain cortical volume, total left and right hemisphere cortical volume, left and right cerebral-white-matter, subcortical, supratentorial, and intracranial volume; (Bs> 0.030, p_fdrs_<0.0021). All associations were significantly larger for the cognitive PGS relative to the non-cognitive PGS (all p_diff_<0.05).

#### Global White Matter Tracts (Figure 4A**; Supplemental Table 6).**

The non-cognitive PGS was positively associated with the global measure of mean diffusivity (B=0.029, p_fdr_=0.023). The cognitive PGS was not associated with any global atlas tract fiber measures, but the associations for the non-cognitive PGS were not significantly stronger than that observed for the cognitive PGS.

#### Volume (Figure 4B; Supplemental Table 7)

The cognitive PGS was positively associated with the following 13 regional brain volumes: (7 lateral and 3 bilateral: 1-2) bilateral inferior temporal gyri, 3-4) bilateral precentral gyri, 5-6) bilateral superior temporal gyri, 7) right (R) banks of superior temporal sulcus, 8) R caudal anterior cingulate, 9) R inferior parietal gyrus, 10) R middle temporal gyrus, 11) R pars orbitalis, 12) R temporal pole, 13) left (L) rostral anterior cingulate (B=0.032-0.0383, p_fdr_=0.005-0.035). The following regional associations were significantly greater for the cognitive PGS relative to the non-cognitive PGS: rh-pars orbitalis, lh-superiortemporal, rh-middle temporal, rh-inferior parietal, and bilateral inferior temporal volumes. The non-cognitive PGS was not signficantly associated with any regional cortical volume.

#### DTI-FA (Figure 4C; Supplemental Table 7)

The cognitive and non-cognitive PGS were associated with 1 bilateral white matter tract and 10 white matter tracts (1 unilateral, 1, interhemispheric, and 4 bilateral), respectively. The cognitive PGS was positively associated with the bilateral corticospinal/pyramidal tract (B=0.035-0.041, p_fdr_=0.001-0.005). Tracts associated with the non-cognitive PGS included: 1-2) bilateral corticospinal/pyramidal, 3-4) bilateral superior corticostriatal-frontal cortex, 5-6) bilateral superior corticostriatal-parietal cortex, 7-8) bilateral superior corticostriatal, bilateral corpus callosum, and bilateral forceps minor (Bs=-0.029-0.038, p_fdrs_=0.001-0.046). Both the forceps minor and the left superior corticostriatal-frontal cortex showed associations with the non-cognitive PGS that were of significantly greater magnitude (p_diffs_<0.05) than with the cognitive PGS.

## DISCUSSION

Our study of phenotypic correlates of genetic propensity for cognitive and non-cognitive factors during middle childhood/early adolescence (ns=2,968-5,517) revealed four broad findings: *First,* cognitive and non-cognitive polygenic liability both showed associations with cognition scores, socioeconomic status, and psychopathology. Notably, the associations between cognition scores and the cognitive PGS were of significantly greater magnitude than those of the non-cognitive PGS, but there were no other significant differences between the PGSs. *Second,* as revealed by our *within-family* analyses, associations between the cognitive PGS and some outcomes (e.g., cognition, anorexia) were consistent with direct genetic effects and/or evocative/active rGE. *Third*, the cognitive PGS was more strongly associated with greater regional cortical volumes than the non-cognitive PGS, while the non-cognitive PGS generally showed greater magnitude of associations with white matter tract fractional anisotropy. *Fourth*, *within-family* analyses revealed significant within-family associations between the cognitive PGS and right hemisphere caudal anterior cingulate and pars orbitalis, while the non-cognitive PGS had significant within-family associations with the FA tract right superior corticostriate-frontal cortex. Overall, these findings converge with other recent data to suggest that the phenotypic correlates of genetic liability to cognitive and non-cognitive factors underlying educational attainment are present during childhood, prior to the completion of most education, and largely mirror patterns in adults.

### Polygenic Scores for Cognitive and Non-cognitive Factors Related to Educational Attainment Have Shared and Unique Correlates

The cognitive PGS was associated with higher fluid intelligence, crystallized intelligence, and a total combined score. The largest association was with crystallized intelligence, mirroring a prior ABCD study where researchers found that an intelligence PGS was most predictive of the crystallized intelligence score (Loughnan et al., 2023). The cognitive PGS was also associated with lower neighborhood deprivation and higher family income. These findings reflect prior research showing that cognitive PGS are associated with increased family income as well as better neighborhood environment (Judd et al., 2020), and that cognitive development in children is linked to both family income and their grandfather’s occupational status (Najman et al., 2004). Interestingly, we found that the cognitive PGS was associated with delayed discounting and greater risk tolerance, as well as decreased odds for initiating nicotine use; this latter finding is congruent with previous research showing that greater educational attainment may correlate with decreased likelihood of substance initiation (Fothergill et al., 2008; Morin et al., 2012). The cognitive PGS was also associated with decreased odds of an ADHD diagnosis and lower psychotic-like experience (PLE) severity scores, but greater risk for anorexia. This latter finding is generally in line with work showing a positive association between cognitive performance and educational attainment and anorexia (Demange et al., 2023; THE BRAINSTORM CONSORTIUM et al., 2018; Watson et al., 2019), while the correlation with lower severity of PLEs is highly consistent with other work showing PLEs to be related to decreased school performance (Davies et al., 2018; Villa et al., 2023) and general cognitive deficits (e.g., executive functioning; Sheffield et al., 2018).

While the non-cognitive PGS was also associated with higher crystallized and total cognition scores, associations with the non-cognitive PGS were significantly smaller than with the cognitive PGS (p_diffs_ 2.2e-16 - 1.51E-12). Similar to findings with the cognitive PGS, the non-cognitive PGS was associated with higher family income, lower risk of ADHD, and lower PLE severity scores. The association of both PGSs with lower likelihood of an ADHD diagnosis is very consistent with previous studies (de Zeeuw et al., 2014). While some measures were only significantly associated with the cognitive PGS (e.g., risk-taking, nicotine initiation, anorexia), only the cognition scores (crystallized, fluid, and total) showed effect sizes which statistically differed (p_diff_ < 0.05) for the cognitive PGS vs. non-cognitive PGS. In contrast to the phenotypic outcomes described above, neuroimaging measures showed a greater number of associations which significantly differed for the cognitive and non-cognitive PGSs.

Globally, all volume measures (N = 9; Figure 4) were significantly associated with both the cognitive and non-cognitive PGSs, but all associations were significantly stronger for the cognitive PGS. In contrast, there were no significant differences between the effect sizes of the two PGSs across the assessed global diffusion tensor imaging modalities. While mean diffusivity was only significantly associated with the non-cognitive PGS, this effect size did not significantly differ from that of the cognitive PGS. The cognitive PGS was associated primarily with regional cortical volumes (N = 13; Figure 4), while the non-cognitive PGS was associated only with regional white matter modalities (N =10; Figure 4). Of note, across the majority of assessed non-global metrics, there were significant differences in the strength of association between the two PGS. For example, bilateral inferior temporal volume, a region implicated in studies of academic achievement, showed a significantly stronger association with the cognitive PGS (Mackey et al., 2015). Conversely, the tract forceps minor, which has previously been implicated in executive functioning and achievement (Loe et al., 2019), showed a significantly stronger (negative) association with the non-cognitive PGS. Overall, these findings suggest that cognitive skills that contribute to educational attainment may relate more strongly to cortical volumes, while non-cognitive factors of educational success may be better reflected in the micro structure of the white matter tracts connecting between brain regions.

**Figure 4.**
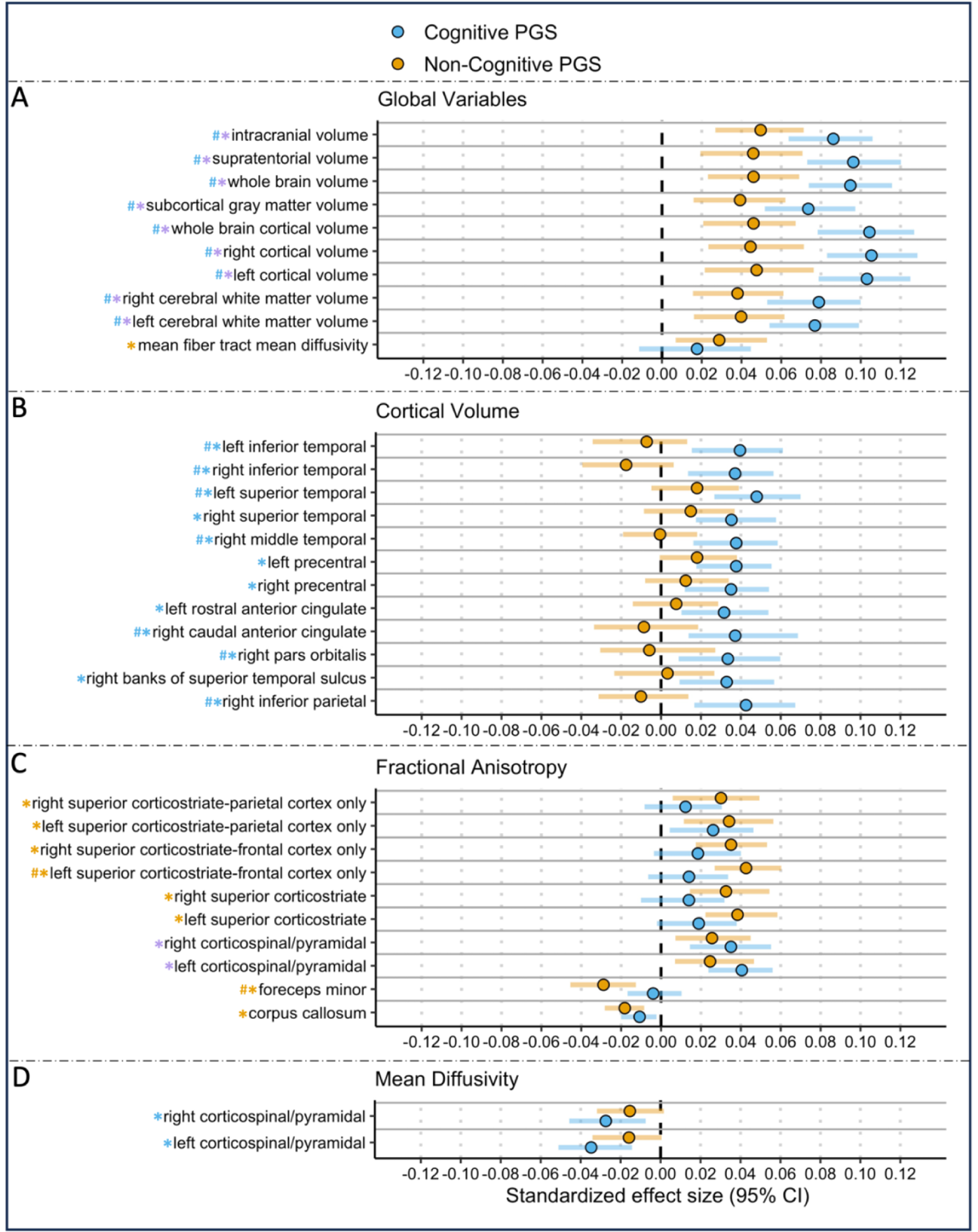
Significant Associations Between Cognitive and Non-cognitive PGS and Neural Indices of Interest. Significant associations between cognitive and non-cognitive PGS and significant imaging Gorelik 14 modalities including: A) global brain indices, B) cortical volume, C) fractional anisotropy, and D) mean diffusivity. Blue, orange, and purple asterisks correspond to significant associations (p_fdr_<0.05) between the outcome measure and cognitive, non-cognitive, or both PGS respectively. Blue hashtags correspond to associations with cognitive PGS that are of significantly greater magnitude than for the non-cognitive PGS, while orange hashtags represent the opposite.

**Figure 5:**
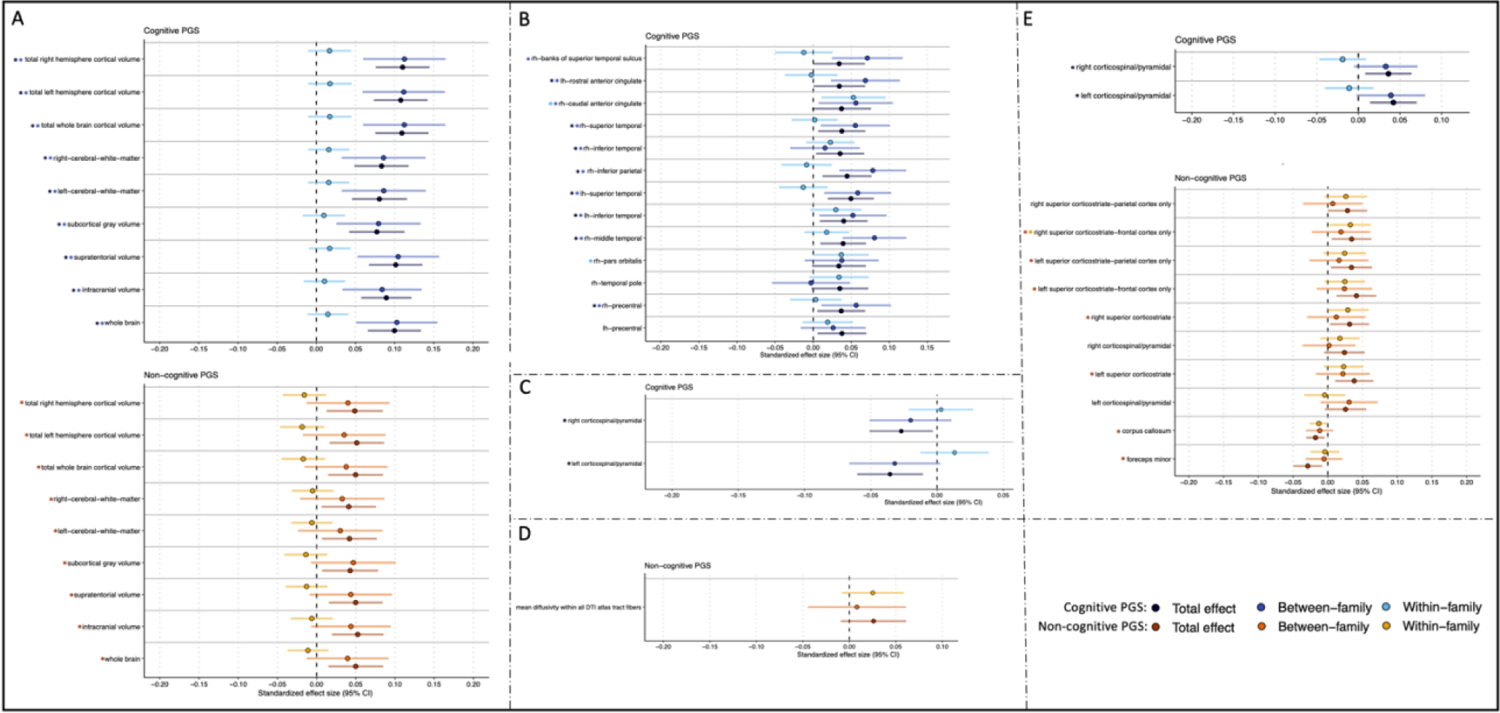
Total, Between, and Within-family Estimates for the Associations Between Cognitive and Non-cognitive PGS and Neural Indices of Interest. Total, Within- and between-family associations between Cognitive and Noncognitive PGS (p <0.05) and significant measures for the neural indices of A) global volume, B) regional volume, C) regional MD, D) average MD, and E) regional FA. (i.e., outcomes with p_fdr_<0.05 in **Figure 4 and Supplemental Table 8**). For the cognitive PGS, black, dark blue, and light blue asterisks correspond to significant total, between-, and within-family associations, respectively. For the non-cognitive PGS, red, orange, and yellow asterisks correspond to significant total, between-, and within-family associations, respectively.

### Evidence for Direct and Indirect Genetic Effects on Phenotypes

Using *within-sibling* analyses to contrast within-vs. between-family estimates for phenotypic correlates, we sought to determine whether associations with the polygenic scores were consistent with possible confounding by passive rGE or other mechanisms (e.g., assortative mating). The cognitive PGS showed significant *within-sibling* associations with all three cognition measures (fluid, crystallized, and total score; **Supplemental Table 5,** Figure 3), suggesting that associations between the cognitive PGS and these cognitive measures reflect direct genetic effects and/or evocative/active rGE, rather than passive rGE or other confounding factors. Interestingly, the cognitive PGS showed a significant *within-sibling* association with anorexia, suggesting direct genetic effects or evocative/active rGE influence this relationship as well. The cognitive PGS also had significant *within-sibling* associations with the right hemisphere caudal anterior cingulate and pars orbitalis measures, both of which have been associated with the salience brain network and cognitive salience (Snyder et al., 2021). No other *within-sibling* associations were significant, consistent with effects of passive rGE or other confounding factors that vary between families (de Zeeuw et al., 2014; Mitchell et al., 2022).

The non-cognitive PGS showed a significant *within-sibling* association with FA tract right superior corticostriate-frontal cortex, which is associated with many relevant pathways, including learning and reward sensitivity (Shipp, 2017). The null *within-sibling* findings for all other phenotypes for the non-cognitive PGS suggest that associations between the non-cognitive PGS and cognition scores and other outcomes may be due to passive rGE (e.g., parents with greater genetic predisposition for motivation or curiosity might enroll their children in additional courses or training) or confounding factors that vary between families, such as assortative mating. Further, these null findings do not seem to be due solely to the lower sample size of these models (N=1,702), as the non-cognitive PGS was associated with crystallized and total neurocognition scores when we randomly resampled 1,702 individuals 5,000 times and averaged across regression models in these smaller samples, and these “total” effect sizes were similar to the full effects presented in Figures 2 **and 4** (see “Total” effects in **Supplemental Tables 5-6**). Our findings for the non-cognitive PGS contrast with a recent preprint from Malanchini et al., 2023 who found evidence that both passive *and* evocative/active rGE mechanisms appear to contribute to the association between non-cognitive genetic factors and academic achievement in a sample of adolescents (Malanchini et al., 2023). Our conflicting results may be partly explained by a combination of social, educational, and cultural differences between the participants of the ABCD study (American) and the Malanchini study (England and Wales), which may influence related non-cognitive factors of educational attainment (Breinholt & Jaeger, 2020; Mendez & Zamarro, 2018).

### Findings in Middle Childhood and Early Adolescence Largely Mirror Findings in Adult Samples

The majority of the current findings in this early adolescent sample mirror findings in Demange et al., 2021 as well as other majority adult samples (Mitchell et al., 2022). Similar to Demange et al., the cognitive PGS was associated with higher fluid intelligence, crystallized intelligence, and a total combined score, while the non-cognitive PGS was only associated with higher crystallized and total scores. Additionally, our findings that both the cognitive and non-cognitive PGSs were associated with measures related to socioeconomic status (Carter et al., 2019) are congruent with findings from Demange et al., and others (Carter et al., 2019). Further, our results for the assessed neuroimaging phenotypes largely mirror those of the Demange et al. study, as well as prior research on academic achievement which has found that larger global brain volumes were positively associated with polygenic scores for educational attainment in a sample of young adults (Demange et al., 2021; Mitchell et al., 2020). Given the congruent findings between previous studies performed in samples of adults (de Zeeuw et al., 2014; Demange et al., 2021) it may be that neural mechanisms associated with academic achievement are developed as early as middle childhood and may be temporally stable until later in life (Lövdén et al., 2020).

However, there were several domains where findings between adolescent and adult samples partly diverge. Notably, Demange et al., 2021 found that their cognitive factor was genetically correlated with *lower* risk tolerance, whereas we observed a *positive* association between the cognitive PGS and risk-taking in this sample. These opposite findings may potentially be explained by age differences across the samples: for example, in adolescents, cognitive abilities have previously been linked to greater risk-taking (Mischel et al., 1989). Another potential reason for this divergence is that adolescence is a neurodevelopmental period characterized by delayed cognitive control development (Shulman et al., 2016). As such, individual differences in cognitive control will be more tightly linked to risk-taking in adolescence than in adulthood. Additionally, Demange et al., found negative genetic correlations between the non-cognitive factor and health-risk behaviors, whereas the current study found no significant associations between the non-cognitive PGS and risk tolerance, decision-making, or substance initiation. However, our null findings may be due to a combination of low endorsement of risky behaviors in our younger adolescent sample, as well as limitations of these measures in ABCD (i.e., the ABCD study has fewer assessments of health-risk behaviors). In the domain of psychopathology, we found that both the cognitive and non-cognitive PGSs were *negatively* associated with PLE severity scores, diverging from Demange et al.’s observed *positive* relationship between the non-cognitive factor and schizophrenia risk, as well as other studies that have linked greater creativity (Rajagopal et al., 2023) and other aspects of academic success to greater risk for schizophrenia (Karlsson, 2004). The negative associations we observed between the PLE severity scores and both PGSs may suggest that cognitive and non-cognitive factors capture aspects of psychosocial functioning that are protective against development of prodromal psychosis in this adolescent sample (Junghoon et al., 2023).

### Strengths and Limitations

Strengths of this study include a relatively large sample size, assessments taken during a developmentally important time period (adolescence), deep phenotyping across multiple psychosocial and neuroimaging measures, and the incorporation of within-family analyses to control for potential confounding factors. However, this manuscript is not without limitations. First, this is a cross-sectional study in a European ancestry subsample of individuals whose caregivers volunteered to participate in the research study (Schoeler et al., 2023). This limits study generalizability, which may be especially important for better understanding potential relationships between facets of academic achievement and factors related to health and well being. Second, the outcome measures studied were constrained by what was available in the ABCD Study, and therefore do not include all potential measures of interest (e.g., personality traits that are considered to be important to non-cognitive aspects of educational attainment and success; Humphries & Kosse, 2017; Malanchini et al., 2023; Tucker-Drob et al., 2016; von Stumm et al., 2011). Relatedly, we were unable to assess all of the same imaging modalities that prior papers have reported on, including mode of anisotropy. Third, it is still unclear what exactly is represented by the “non-cognitive” factor GWAS used to create polygenic scores for the current study, and whether this non-cognitive factor would differ if the input GWASs were derived from samples of different ages (e.g., childhood, adolescence, older age). The authors of the original study (Demange et al., 2021) posit that preferences of risk and decision-making, personality traits, and socially desirable behaviors contribute to the non-cognitive factor of educational attainment captured by their study, as evidenced by patterns of genetic correlations with relevant phenotypes. However, as the authors note, the original GWAS of cognitive performance (Lee et al., 2018) may not have captured *all* relevant aspects of cognitive ability across the lifespan, and thus the separation of “cognitive” vs. “non-cognitive” may be incomplete. Future studies should focus on explicating the nature of the non-cognitive factors and traits that contribute to academic achievement.

## Conclusion

Overall, the results of this study provide evidence that as early as adolescence, polygenic scores for cognitive and non-cognitive facets of educational attainment share both overlapping and unique associations with psychosocial outcomes and neuroimaging measures. We speculate that the majority of these PGS associations are stable across adolescence and adulthood; however, further studies are needed before such a conclusion can be made. As the participants of the ABCD Study continue to age, this will be an invaluable sample in which to characterize the degree to which genetic and environmental effects on academic achievement, psychopathology, health behaviors, and neural phenotypes change across development into adulthood.

## Acknowledgements

We are thankful to families who have participated in the ABCD Study as well as study staff and investigators.

## Declarations

### Ethics Approval

Working with ABCD NDA data was approved by the Washington University in St. Louis Institutional Review Board: IRB ID#201708123.

### Data availability

All ABCD data used in this study are available through the National Institute of Mental Health Data Archive (NDA), which may be accessed here (https://nda.nih.gov).

### Code availability

https://github.com/WashU-BG/ABCD_cog_non_cog_2023

### Author Contributions

ECJ, AJG, NRK, WZ, JB, APM, SEP, and SL conceptualized the study and planned analyses. AJG, SEP, APM, and NRK curated the phenotypic data. SL and ECJ created the polygenic scores, and AJG, SL, ASH, and ECJ performed statistical analyses. PA, DAAB, AJG, and ECJ contributed to data visualizations. AJG, ECJ, RB, SL, JB, NM, and HM drafted the initial manuscript, and all coauthors edited the manuscript and provided important intellectual feedback.

