## supplemental methods for "Associations Between Polygenic Scores for Cognitive and Non-cognitive Factors of Educational Attainment and Measures of Behavior, Psychopathology, and Neuroimaging in the Adolescent Brain Cognitive Development Study"

**Neuroimaging Acquisition and Processing for Imaging Phenotypes**

Imaging data from the baseline assessment of ABCD of individuals from European descent are available for 5556 out of 5560 participants, out of the 11,556 usable brain scans. More details on the description of the imaging acquisition procedures, processing, and analysis methodology in ABCD can be found elsewhere (Casey et al., 2018; Hagler et al., 2019). Briefly, 1mm isotropic T1-weighted structural images were obtained via 3T MRI scanners (Siemens, Phillips, and GE) using either a 32- or 64-channel head-and-neck coil and completed T1-weighted and T2-weighted structural scans (1mm isotropic). MRI scan protocols were harmonized across the three MRI vendor platforms to minimize variability. Real-time motion detection and correction was implemented to mitigate the influence of head motion. Hagler et al. (2019) provides a comprehensive description of the quality-control measures conducted on the processed imaging data. Here we provide a brief summary for each imaging modality used:

*Structural MRI Processing*

For structural MRI (sMRI) metrics, structural neuroimaging processing was completed using FreeSurfer version 5.3.0 through standardized processing pipelines (Hagler et al., 2019). Participants that did not pass FreeSurfer Quality Control measure (i.e., at least one T1 scan that passed all quality control metrics) were excluded from analyses (n= 69). A detailed description of the quality-control measures conducted on the processed data is provided in Hagler et al. MRI analyses included only participants whose structural MRI reconstructions passed QC tests. Cortical reconstruction and volumetric segmentation were performed by the ABCD Study® Data Acquisition and Integration Core using the FreeSurfer image analysis suite (<http://surfer.nmr.mgh.harvard.edu/>). This pre-processing includes removal of non-brain tissue using a hybrid watershed/surface deformation procedure (Ségonne et al., 2004), automated Talairach transformation, segmentation of the subcortical white matter and deep gray matter volumetric structures, intensity normalization, tessellation of the gray/white matter boundary, automated topology correction, and surface deformation following intensity gradients (Hagler et al., 2019). Images were registered to the Desikan atlas, which was based on individual cortical folding patterns to match cortical geometry across subjects. The cerebral cortex was parcellated into 34 regions per hemisphere based on the gyral and sulcal structure. For the imaging analyses, cortical gray matter volume and total cerebral white matter volume aligned to the Desikan atlas were extracted from ABCD data release 3.0. nine global metrics were analyzed first and included total intracranial and whole brain volumes; total and bilateral cortical volume; total supratentorial volume; total subcortical gray matter volume; and bilateral cerebral white matter volume. Only sMRI data that passed these QC tests (n=5,513) was retained.

*Diffusion Weighted Imaging Processing*

diffusion MRI (dMRI) metrics included mean diffusivity (MD) and fractional anisotropy (FA) metrics. dMRI metrics were calculated with a linear estimation approach with log-transformed diffusion-weighted signals (Basser et al., 1994). To create MD and FA metrics, first, tensor matrices were diagonalized using singular value decomposition, resulting in three eigenvectors and three corresponding eigenvalues. FA is derived from the eigenvalues (Basser & Pierpaoli, 1996). MD is derived from the mean of the eigenvalues. Major white matter tracts are labeled using AtlasTrack, a probabilistic atlas-based method for automated segmentation of white matter fiber tracts; (Hagler et al., 2009)^6^ 16 tracks were used in our analysis. All analyses including dMRI metrics included mean motion, fmax and fmin as additional covariates. Only dMRI data that passed QC tests (n=5,258) was retained.

**Genetics**

*Genotyping QC, Imputation, and Polygenic Scores*

Quality control on genomic data (data release 3.0; n= 11,099) was performed using default Rapid Imputation and COmputational PIpeLIne for Genome-Wide Association Studies (RICOPILI; (Lam et al., 2020))^7^ settings. Of the 10,585 individuals who passed initial QC checks, 6,787 parents/caregivers indicated that their child’s race was only “white,” and 5,561 of those individuals did not endorse any Hispanic ethnicity/origin. Using data from unrelated individuals (pi-hat ≤ 0.2) and an LD pruned set of common (MAF>0.05) and non-palindromic SNPs (and excluding the MHC region and chromosome eight inversion), principal components analysis (PCA) was performed in RICOPILI using EIGENSTRAT (Price et al., 2006) to confirm the genetic ancestry of these individuals. This was done by merging the ABCD Study data with the 1000 Genomes reference panel (1000 Genomes Project Consortium et al., 2015), computing the mean and standard deviations for the 1000 Genomes European ancestry population for the top 6 PCs, and establishing that the previously identified group of 5,561 participants were genetically similar to the 1000 Genomes European panel by assessing whether they fell within 3 SDs of the mean for the top 6 PCs within this 1000 Genomes European reference population. After another round of QC on this subset, 5,556 European ancestry individuals were retained. Only individuals of European ancestry were analyzed as the discovery GWAS only included individuals of European ancestry and there is poor predictive utility across ancestries which may lead to erroneous conclusions (e.g., false negatives; Martin et al., 2019).

The QCed genotype data were then imputed to the TOPMed imputation reference panel (Taliun et al., 2021). Imputed dosages were converted to best-guess hard-called genotypes, and single nucleotide polymorphisms (SNPs) with Rsq > 0.8 and MAF > 0.01 were retained for PGS analyses. For the PGS of both cognitive and non-cognitive PGS, the number of MCMC iterations was set at 10,000, and the number of burn-ins was set at 5,000. After deriving SNP weights using PRS-CS, we then used PLINK 1.9’s (Chang et al., 2015) --score command to produce PGS in the ABCD sample.

*Within Sibling Analyses:*

For the *within sibling* analyses, we included both the family mean PGS and a sibling’s deviation from their family mean PGS as predictors in a mixed-effect model, as has been done previously (Selzam et al., 2019):

$$Y_{ijk} =\beta_{W}({PGS}_{ij} -\underline{{PGS}_{j}}) +\beta_{B}(\underline{{PGS}_{j}}) +\gamma_{j}+\gamma_{k}+\varepsilon_{ijk}$$

where $Y_{ijk}$represents the outcome for sibling *i* in family *j* at site *k*, ${PGS}_{ij}$ represents the polygenic score for sibling *i* in family *j*, $\underline{{PGS}_{j}}$ represents the mean PGS for family *j*, $\gamma_{j}$ represents a random intercept for family *j*, $\gamma_{k}$ represents a random intercept for site *k*, and $\varepsilon_{ijk}$ represents the independent random error for each individual *i* in family *j* at site *k*. We also included the same fixed-effect covariates (age, sex, and genetic principal components) as in our primary models described above (not shown in equation). In this way, we were able to partial the variance in the outcome explained by the PGS into between-family ($\beta_{B}$) and within-family ($\beta_{W}$) effects. Because the sample of siblings (N = 1,702) was considerably smaller than the full analytic sample (N = 5,517), we derived the population-level estimates (the “Total” effects) between PGS and outcomes in a sample of equivalent size (and statistical power). To this end, we randomly sampled 1,702 individuals from the full analytic sample and ran the primary regression models in this smaller sample. We repeated this re-sampling procedure 5,000 times, ensuring that the numbers of cases and controls for binary outcomes were proportionate to that of the full sample, and then calculated the average beta across these 5,000 samples. We calculated the standard deviation of the sampling distribution of betas (equivalent to the standard error of the mean), a z-score (mean beta/SE), and a p-value of the z-score. For these post-hoc analyses, we considered associations with p < 0.05 to be significant and present uncorrected p-values in **Supplemental Table 5-6**.

**Statistical analyses**

Analyses were performed using R Statistical Software version 4.3 (R Core Team 2021). Associations between PGS and outcomes were estimated using mixed effects models (lmer() for continuous and glmer() for dichotomous outcomes) in the lme4 R software package version 1.1 to account for non-random clustering of data (Austin, 2010; Bates et al., 2015). An additional warning statement and the bobyqa optimizer was used in case models did not converge on a local minima with first attempt. PGS were normalized to z scores (i.e., mean = 0, standard deviation=1) before including them as simultaneous predictors in regressions. Age, sex, and the first 10 genetic principal components were included as fixed effect covariates, and random intercepts were included for family ID and recruitment site. For imaging models, recruitment site was replaced by MRI serial number. Imaging models also included MRI manufacturer, global brain metrics representing the mean for each modality, and mean motion for DTI as fixed effects. We used false discovery rate (FDR; Benjamini & Hochberg, 1995) to account for multiple testing (p_fdr_<0.05); FDR was applied separately to the non-imaging phenotypes and each respective imaging modality.

We deviated from our pre-registered analyses in two ways: First, we tested whether the regression coefficients for the cognitive and non-cognitive PGS significantly differed from each other in each model (p_diff_). To do this, we used the ‘linearHypothesis()’ function from the ‘car’ package in R (Weisberg, 2019) to compare a restricted model with the full model, and a significant chi-square test statistic indicates a significant difference in the regression coefficients for the cognitive PGS vs. the non-cognitive PGS. We considered p < 0.05 to be significant.

Second, we used *within-sibling* analyses to assess whether within-sibling variation in cognitive and non-cognitive PGS are associated with outcomes that were significantly associated with PGS in our primary analyses (aside from familial SES due to sibling similarity). These within-sibling analyses provide additional inference into the the potential direct PGS effects on outcomes as any significant associations would be unconfounded by population stratification, assortative mating, passive gene-environment correlations, and other potential confounders that can plague population-level estimates (though it should be noted that active and evocative rGE will still influence within-sibling variation in PGS effects; Brumpton et al., 2020; Howe et al., 2022; Young et al., 2018). For these analyses, we included both the family mean PGS and a sibling’s deviation from their family mean PGS as predictors in a mixed-effect model, as has been done previously (Selzam et al., 2019). Because the sample of siblings (N=1,702) was considerably smaller than the full analytic sample (N=5,517), we derived the population-level estimates (the “Total” effects) between PGS and outcomes in a sample of equivalent size (and statistical power). To this end, we randomly sampled 1,702 individuals from the full analytic sample and ran the primary regression models in this smaller sample. We repeated this re-sampling procedure 5,000 times, ensuring that the numbers of cases and controls for binary outcomes were proportionate to that of the full sample, and then calculated the average beta across these 5,000 samples. For these post-hoc analyses, we considered associations with p < 0.05 to be significant and present uncorrected p-values in **Supplemental Table 5-6**.
